# Co-infection with SARS-COV-2 Omicron and Delta Variants Revealed by Genomic Surveillance

**DOI:** 10.1101/2022.02.13.22270755

**Authors:** Rebecca J Rockett, Jenny Draper, Mailie Gall, Eby M Sim, Alicia Arnott, Jessica E Agius, Jessica Johnson-Mackinnon, Elena Martinez, Alexander P Drew, Clement Lee, Christine Ngo, Marc Ramsperger, Andrew N Ginn, Qinning Wang, Michael Fennell, Danny Ko, Linda Huston, Lukas Kairaitis, Edward C Holmes, Matthew N O’Sullivan, Sharon C-A Chen, Jen Kok, Dominic E Dwyer, Vitali Sintchenko

## Abstract

We identified the co-infection of the SARS-CoV-2 Omicron and Delta variants in two epidemiologically unrelated patients with chronic kidney disease requiring haemodialysis. Both SARS-CoV-2 variants were co-circulating locally at the time of detection. Amplicon- and probe-based sequencing using short- and long-read technologies identified and quantified Omicron and Delta subpopulations in respiratory samples from the two patients. These findings highlight the importance of genomic surveillance in vulnerable populations.

## MAIN

Since the declaration of the COVID-19 pandemic by the World Health Organization (WHO) on March 11^th^ 2020, SARS-CoV-2 has gradually evolved into phylogenetically distinct lineages, some of which have been designated Variants of Concerns (VOCs).^1,2^ These variants differ in terms of transmissibility, capacity to cause severe disease and the ability to evade post-vaccination derived immunity. The global prevalence of individual VOCs in different global regions has been affected by the timing and location of their emergence and the corresponding measures for COVID-19 control measures.^3^ Development and implementation of viral genomic surveillance and rapid sharing of genomic data has provided a critical capacity to distinguish and monitor SARS-CoV-2 variants and conduct risk assessments of their significance. Co-infection with different SARS-CoV-2 lineages was rarely reported during the first COVID-19 wave in 2020 prior to the introduction of vaccination programs,^4–6^ but it has been suggested that such co-infections could lead to greater severity and disease duration.^6^ However, co-infections involving either VOC Delta or VOC Omicron have not yet been reported, nor have they been reported in immunosuppressed hosts, which may drive saltational evolution.^7^ Here, we report the first cases of co-infection with Delta and Omicron in two immunocompromised individuals at risk of severe COVID-19 disease identified during local co-circulation of both SARS-CoV-2 lineages.

Case A was a patient aged between 60 - 70 years who returned a positive SARS-CoV-2-specific polymerase chain reaction (PCR) result from a nasopharyngeal swab after presenting to the Emergency Department of a Sydney hospital with mild respiratory symptoms. Case B was a patient, aged between 50-60 years diagnosed by SARS-CoV-2 PCR after presenting to the same hospital with fever. Samples from both cases underwent whole genome sequencing as part of the prospective genomic surveillance program in New South Wales (NSW), Australia.^8^ Only genomes confidently assigned to SARS-CoV-2 lineages (Supplementary Figure S1) are reported to the health authorities and shared globally via GISAID (https://www.gisaid.org). In contrast to the majority of community samples sequenced, those obtained from Cases A and B had unexpectedly high numbers of “heterozygous” (i.e., mixed nucleotides at a single site) calls (Supplementary Figure S2) and could not be unambiguously assigned to a SARS-CoV-2 lineage by the Pangolin software. This observation triggered a case review which revealed that both patients had chronic kidney disease due to type 2 diabetes, obesity and ischaemic heart disease. In addition, both were receiving haemodialysis treatment for 4-5 hours thrice weekly at the same community dialysis centre and therefore potentially exposed to multiple COVID-19 cases during treatment session. Given the high community incidence of COVID-19, infection control measures implemented at the dialysis centre to prevent nosocomial transmission included physical distancing and masking of patients at all times, decontamination of treatment stations and dialysis equipment after each session, four-point personal protective equipment use by all clinical staff and patient surveillance testing by PCR at the time of each treatment. Despite the similarities in patient demographics, they were unknown to each other, had not received haemodialysis at the same time nor used the same equipment or treatment station.

Neither patient had prior evidence of COVID-19 infection. Case A had received two doses of the COMIRNATY® (Pfizer) vaccine with the second dose ten weeks prior to diagnosis. Case B remained unvaccinated by choice.

PCR did not detect human influenza viruses A or B, respiratory syncytial virus, parainfluenza viruses 1, 2, and 3, human metapneumovirus or rhinovirus in samples from both cases. A sample from an epidemiologically linked household contact of Case A, Case C, who was diagnosed several days after Case A was also sequenced.

Following the observation of high numbers of heterozygous calls in samples collected from Cases A and B, (both exposed to COVID-19 patients potentially infected with different co-circulating lineages), additional viral sequencing and viral culture was performed. Due to the low viral load in the Day 0 samples from both cases, they were only able to be sequenced using Midnight primers and Illumina sequencing, while two longitudinal samples for each case, with increased viral loads were subjected to further analyses (Supplementary Table S1). Two respiratory swabs collected from Case A on Days 2 and 3, as well as two respiratory swabs from Case B collected on Days 3 and 11, were subjected to nucleic acid extraction, quantitative SARS-CoV-2 PCR and genome sequencing using short-read (NextSeq 500 (Illumina)) and long-read (GridION (Oxford Nanopore Technologies; ONT)) protocols using Midnight primers. We also employed the probe-based Illumina Respiratory Viral Oligo panel (RVOP)^9^ to collect reads unbiased by SARS-CoV-2 PCR amplification (see Online Methods for details). Viral yield in samples was variable but still significant and suggesting the presence of viable virus (Table 1). Careful review of the relative frequency of 17 Omicron lineage-defining markers and 10 VOC Delta lineage-defining markers^10^ clearly demonstrated co-infection with both lineages (Figures 1 & 2A). The overall proportion of Delta and Omicron was highly concordant between all three sequencing methods (Figure S3), however four lineage markers showed evidence of amplification bias when SARS-CoV-2 was amplified using Midnight primers (Figure S5). Population analysis of genomic data generated using RVOP methods estimated that the VOC proportions in samples from Case A were 21% Omicron and 77% Delta on Day 2, compared to 45% Omicron and 53% Delta on Day 3. Samples from Case B contained 42% Omicron and 53% Delta on Day 3, and 11% Omicron and 84% Delta on Day 11 (Figures 1, S3-S5). Despite the same pattern of mixed infection, the two cases were not genomically linked in a transmission pathway (Figure 2B). The two Omicron sequences were distinct representatives of the Omicron (sub-lineage BA.1) strain currently predominating in Sydney, while the two Delta sequences belonged to different genomic clusters of Delta (sub-lineage AY.39.1) also circulating locally (Figure 2, Table 1). These conclusions were supported by matching the Omicron sequence from Case A to the Omicron genome recovered from their household contact (Case C, Figure 2).

**Table 1.** SARS-CoV-2 yield in Cases A and B

| Samples |  | SARS-CoV-2 RT-PCR Ct value | SARS-CoV-2 viral load |  | Lineage and NSW cluster designations inferred from genomic data |
| --- | --- | --- | --- | --- | --- |
| | | | cpy/μL | Log<sub>10</sub> | |
| Case A | Day 2 | 17.33 | 98,130,334 | 8.0 | Omicron B.1.1.529 (NSW171);<br>Delta AY.39.1 (NSW130.134.11) |
|  | Day 3 | 23.44 | 404,527 | 5.6 | Omicron B.1.1.529 (NSW171);<br>Delta AY.39.1 (NSW130.134.11) |
|  | Day 3 Culture | 15.37 | 571,255,772 | 8.8 | Delta AY.39.1 (NSW130.134.11) |
| Case B | Day 0 | 31.68 | 246 | 2.4 | Delta AY.39.1 (NSW130.34.128) |
|  | Day 3 | 19.26 | 17,317,514 | 7.4 | Omicron B.1.1.529 (NSW171)<br>Delta AY.39.1 (NSW130.34.128) |
|  | Day 11 | 24.05 | 233,804 | 5.4 |  |

**Figure 1.**
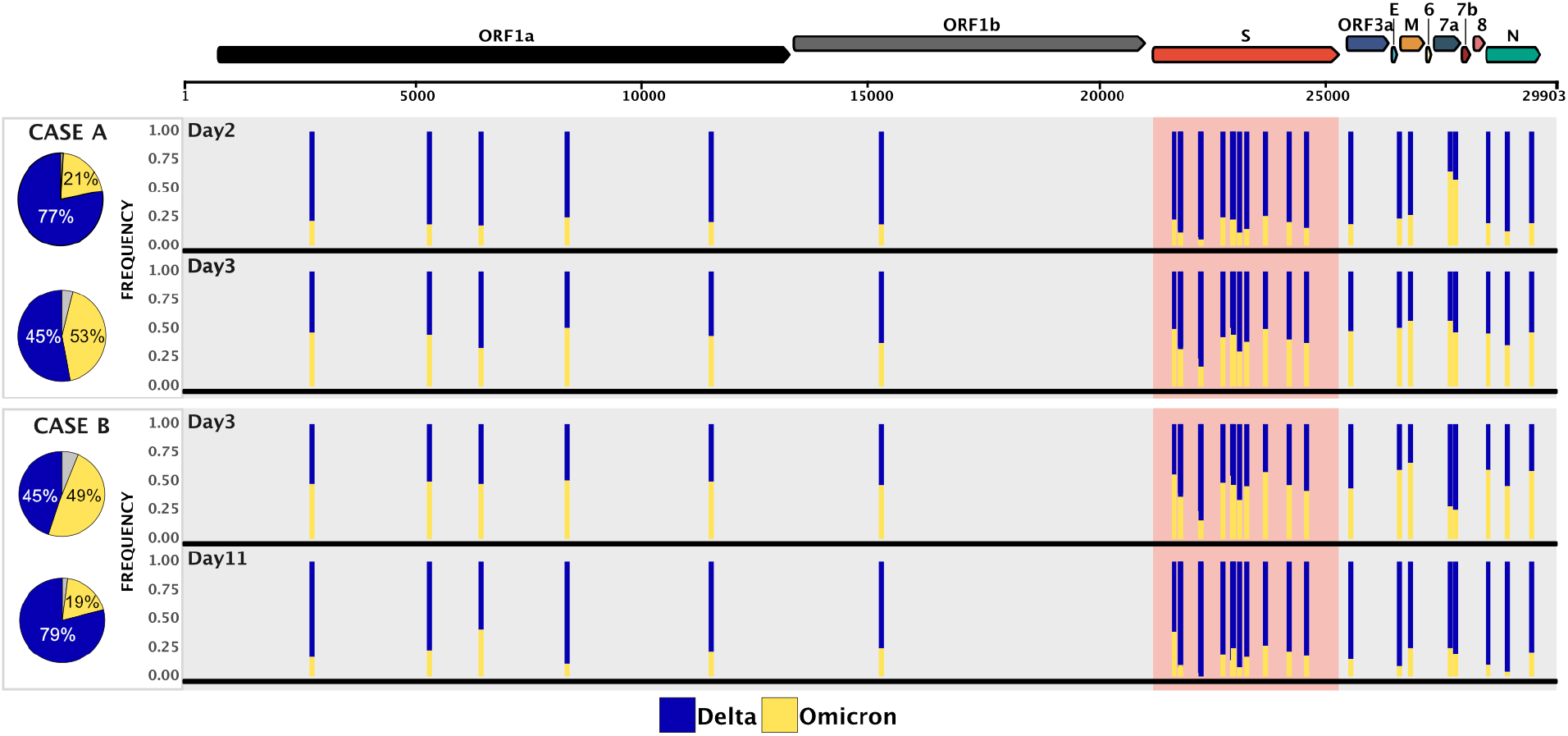
Genome-wide view of the variant frequency of the Delta and Omicron SARS-CoV-2 lineage defining polymorphisms in specimens sequenced using the RVOP SARS-CoV-2 enrichment protocol Pie graphs depict the average population frequency of Omicron and Delta lineage-defining mutations in four clinical samples collected from Cases A and B. Segments in grey represent differences between the average read frequency of Omicron and Delta markers. A total of 27 polymorphisms defining Delta and Omicron lineages are presented in relation to the annotated SARS-CoV-2 genome. The frequency of sequencing reads encoding each mutation is shown by histograms highlighting the constellation of mutations defining each lineage. Blue bars demonstrate the frequency of mutations defining the Delta lineage and yellow bars the Omicron lineage. Due to the close genomic location of lineage defining mutations in the spike region some bars are overlapping. Read frequencies are collected from RVOP data but are highly concordant between sequencing methods and technologies.

**Figure 2.**
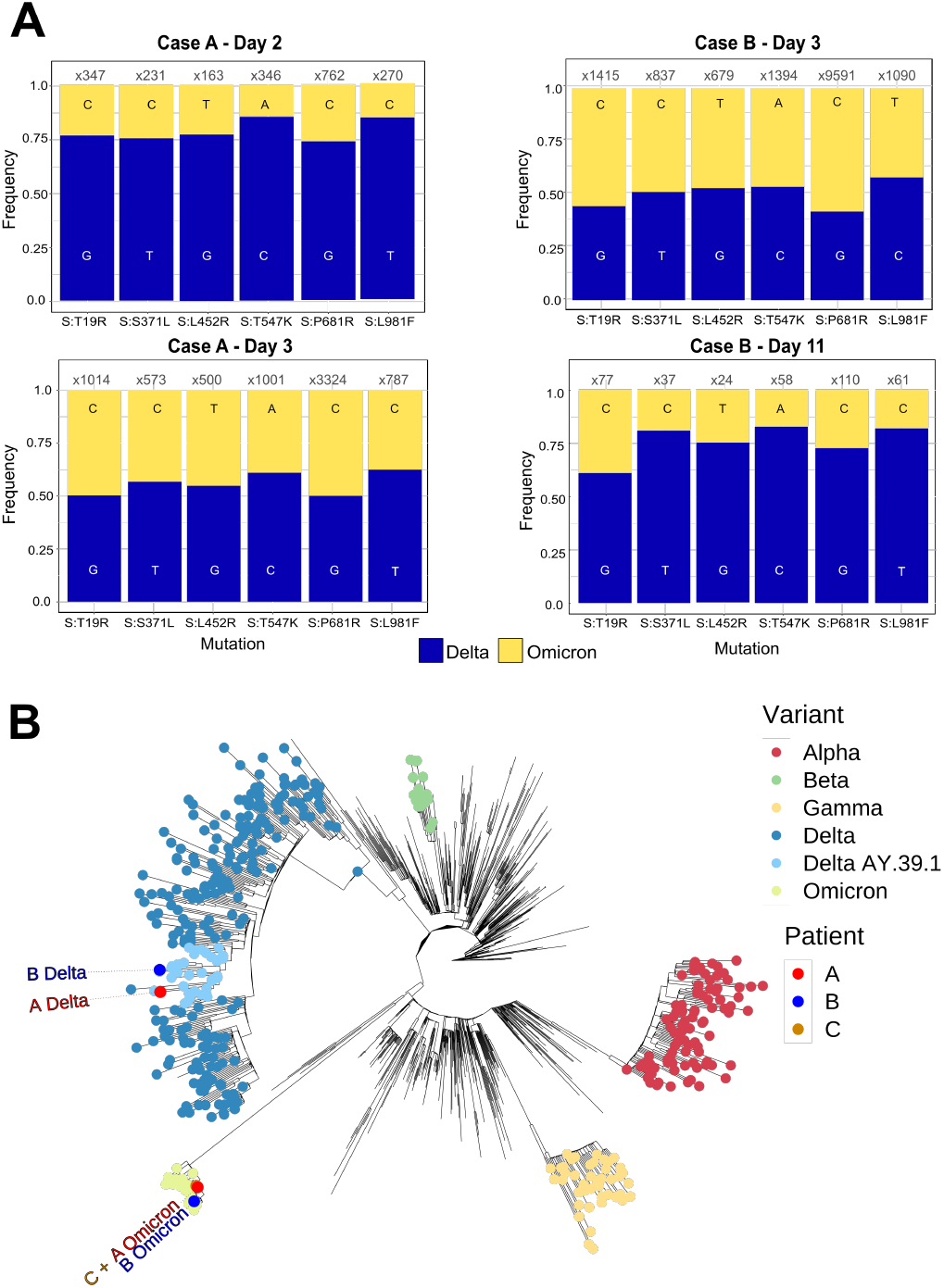
Population and phylogenetic analysis of two cases of SARS-CoV-2 co-infection with Delta and Omicron VOCs. **A**. Population analysis of key lineage defining mutations in the SARS-CoV-2 spike gene for each specimen. Nucleotide frequency and relative coverage of genomic regions specific for either Omicron or Delta. The X-axis represents genomic positions and Y-axis indicates their relative frequencies derived from RVOP data. **B**. Unrooted maximum likelihood phylogeny representing the sequences obtained from Cases A, B and C in the context of global diversity of SARS-CoV-2. Genomes generated as part of this study are labelled individually. The predominant Delta lineage in Australia, AY.39.1, is highlighted. The Delta strains from cases A and B are from separate clades of AY.39.1 circulating in Australia, whereas the two Omicron strains are both in the same sublineage of Omicron (BA.1) which dominated in Australia in December 2021-January 2022. Note that the Omicron samples from patients A and C are identical and hence overlap. Branch lengths are scaled according to the number of nucleotide substitutions per site.

Viral culture of the Case A, Day 3 sample yielded Delta four days post-infection, the consensus genome recovered from this culture and matched the genome reconstructed from the mixed sample. It is likely that Delta had overgrew Omicron as TMPRSS2 enhanced VeroE6 cells are less permissible to Omicron, but highly adapted to Delta infection.^11^ Viral culture was retrospectively and unsuccessfully attempted for the specimen collected from Case B, Day 2. A previously described immunofluorescence assay (IFA)^12^ performed on sera collected from Case 2, Day 3 did not detect SARS-CoV-2 antibodies (i.e. IgG, IgA and IgM IFA titres all <10, trimeric spike IgG negative, nucleoprotein IgG negative).

Although these findings confirm phylogenetically distinct and epidemiologically relevant SARS-CoV-2 variants in both cases, they are not sufficient to conclude whether these cases acquired their dual SARS-CoV-2 co-infection following sequential exposures to individuals with a single lineage infection. Further studies of SARS-CoV-2 within-host population dynamics are required to better understand these processes. The most likely hypothesis for these two cases of SARS-CoV-2 co-infection is the sustained exposure of susceptible, immunosuppressed hosts to multiple patients infected with Delta or Omicron at a time when there was widespread community circulation of both VOCs. Case B appeared to be initially infected by Omicron and superinfected with Delta shortly prior to admission as there were only Omicron sequences in the Day 0 sample and high viral loads of Omicron and Delta obtained from the Day 3 sample.

The identification of phylogenetically distinct and epidemiologically relevant SARS-CoV-2 variants within the same host further expands the relevance of genomic surveillance and highlights the added value of patient and public health context during clinical genomics analysis. The recognition of mixed infections may also affect the selection of appropriate antiviral therapy and infection control measures. Whilst the multiple sequencing methods presented here demonstrated concordant results (Figure S3), not all of them are required to investigate every suspected co-infection case. The RVOP approach was particularly informative as it captured several lineage markers affected by amplification bias due to polymorphisms in whole genome sequencing primers (Supplementary Material, Figures S3-S5). We acknowledge that Cases A and B had relatively equal proportions of sequences representing two lineages in at least one timepoint. The confident identification of minority populations of lineage-specific sequences (e.g., <10%) in more unbalanced populations might be significantly harder; access to longitudinal samples are needed to address these challenges. Genomic epidemiology has rapidly become a high-resolution tool for local and international public health surveillance and disease control. However, the international coverage of SARS-CoV-2 genomic surveillance remains heavily biased towards countries with specialised genomic facilities and research programs.^3,13^ Furthermore, genomic surveillance relies on data sharing by multiple and geographically distributed providers which employ different sequencing and bioinformatic techniques. The reliance on consensus genome data and strict data quality criteria used by genomic laboratories and data sharing environments were designed to minimise the noise from laboratory contamination events and sequencing imperfections. However, such quality metrics can, by design, filter out potentially significant cases associated with high heterozygosity due to mixed viral populations as presented here.

In conclusion, these findings demonstrated the capacity of clinically and epidemiologically informed genomic surveillance to diagnose co-infections with SARS-CoV-2 variants and highlight the needed for deeper analysis of genomic surveillance data in clinical and public health contexts. SARS-CoV-2 co-infections, particularly when they occur in vulnerable hosts may drive saltational evolution, thus emphasising the important role COVID-19 genomic surveillance will play in diagnostic virology, in the era of mass vaccination.

## Data Availability

The SARS-CoV-2 genome sequencing data is available (with human reads removed) in the National Center for Biotechnology Information (NCBI) GenBank (BioProject PRJNA633948).

## ACKNOWLEDGMENTS

The authors acknowledge the Sydney Informatics Hub and the use of the University of Sydney’s high-performance computing cluster, Artemis. The authors are indebted to all researchers and their organisations who have kindly shared SARS-CoV-2 genome data on GISAID.

## AUTHOR CONTRIBUTIONS

Study concept and design by V.S., R.J.R., J.D., M.G., E.S., A.A., and D.E.D. Sample processing and testing by R.J.R., C.L., C.N., M.R., J.A., J.J-M., A.N.G, and Q.W. Sequencing and analysis by R.J.R., E.S., C.L., M.G., J.D., E.M., A.D., V.S. & E.C.H. SARS-CoV-2 viral culture by M.F., D.K., and R.J.R. SARS-CoV-2 serology by L.H. and M.OS. Study coordination by V.S., L.K., S.C-A.C., D.E.D. & J.K. V.S., R.J.R. and A.A. wrote the first manuscript draft with editing from E.C.H., E.S., L.K., S.C-A.C. & D.E.D. The final manuscript was approved by all authors.

## Data availability

The original/raw SARS-CoV-2 genome sequencing data is available in the National Center for Biotechnology Information (NCBI) GenBank (BioProject PRJNA633948).

## Code availability

There were no unique pipelines or source code developed for this project.

