## Supplementary Material for "Co-infection with SARS-COV-2 Omicron and Delta Variants Revealed by Genomic Surveillance"

**METHODS**

***Respiratory virus detection by RT-PCR***. RNA was extracted using either the Viral NA Small volume kit on the MagNA Pure 96 system (Roche Diagnostics GmbH) or RNeasy mini-kit (Qiagen). Both protocols used 200μL of clinical specimen, although minor modifications were performed during the RNeasy extraction, as previously described.^1^ A final elution volume of 100μL was used during MagNA Pure 96 system whereas the RNeasy extraction elute was 30μL.

A previously described RT-PCR^2^, targeting the SARS-CoV-2 nucleocapsid gene was employed to estimate the viral load of clinical specimens from the RNeasy extraction. A commercially available synthetic RNA control (Wuhan-Hu-1 reference sequence, TWIST Biosciences NCBI GenBank accession MN908947.3) was used in 10-fold dilutions starting at 20,000 copies/µL to 2 copies/µL to generate a standard curve and quantify the viral load per microlitre of extracted RNA per specimen (cpy/μL). Additional RT-PCRs were used to investigate the presence of common viral respiratory viruses: human influenza viruses A and B, parainfluenza viruses 1, 2, and 3, respiratory syncytial virus, adenovirus, and rhinovirus^3^

***SARS-CoV-2 culture****.* Respiratory specimens were cultured in Vero E6 cells expressing transmembrane serine protease 2 (VeroE6/TMPRSS2; JCRB1819) as previously outlined.^4^ Briefly, cell cultures were seeded at 1-3x10^4^ cells/cm^2^ in Dulbecco’s minimal essential medium (DMEM, Lonza, Basel, Switzerland) supplemented with 9% foetal bovine serum (FBS, HyClone). Media was replaced within 12 hours with inoculation media containing 1% FBS with the addition of penicillin (10,000 U/mL), streptomycin (10,000 µg/mL) and amphotericin B deoxycholate (25 µg/mL) (Lonza) to prevent microbial overgrowth and then inoculated with 100µL of SARS-CoV-2 RT-PCR positive respiratory sample. The inoculated cultures were incubated at 37˚C in 5% CO_2_ for four days and observed daily for cytopathic effect (CPE). Routine mycoplasma testing using RT-PCR was performed to exclude cell line mycoplasma contamination and culture work was undertaken under physical containment laboratory level 3 (PC3) biosafety conditions. The presence of CPE and increasing viral load as measured by the before mentioned SARS-CoV-2 RT-PCR was indicative of positive SARS-CoV-2 culture. RNA extracts were also subjected to the SARS-CoV-2 genomics workflow as described below.

***Frequency of SARS-CoV-2 variant heterozygosity.*** The frequency of “heterozygosity” (i.e., mixed nucleotides at a single site) during SARS-CoV-2 variant calling is monitored as part of our in-house bioinformatic quality control system, as is the inability to determine a SARS-CoV-2 Pango lineage designation on an otherwise complete and high coverage genome (Figure S1). These two markers signal that a specimen requires repeat extraction, SARS-CoV-2 amplification, library preparation and sequencing.

***SARS-CoV-2 whole genome amplification.*** Tiling PCR was used to amplify the entire SARS-CoV-2 genome from RNA extracts of clinical specimens using primers outlined in the Midnight sequencing protocol.^4^ Each PCR included 12.5µL Q5 High Fidelity 2x Master Mix (New England Biolabs), 1.1µL of either pool 1 or pool 2 10µM primer master mix, 2.5µL of template RNA and molecular grade water was added to generate a total volume of 25µL. Cycling conditions were initial denaturation at 95°C for 2 min, then 35 cycles of: 95°C for 30s, 65°C for 2 min 45s, and a final extension step of 75°C for 10 min. Pool 1 and pool 2 amplicons were combined and purified with a 1:1 ratio of AMPureXP beads (Beckman Coulter) and eluted in 30µL of RNAase free water. Purified products were quantified using Qubit™ 1x dsDNA HS Assay Kit (Thermo Fisher Scientific) and diluted to the desired input concentration for library preparation.

***Amplicon short-read library preparation.*** Purified amplicon pools were used to generate sequencing libraries using Nextera XT (Illumina) according to the manufacturer’s instructions and pooled with the aim of producing 1x10^6^ reads per library. Sequencing libraries were then sequenced with paired-end 76-bp chemistry on the iSeq or MiniSeq or NextSeq (Illumina) platforms.

***Amplicon ONT library preparation.*** In parallel, the purified amplicon pools were also used to generate libraries using SQK-RBK004 (Oxford Nanopore Technologies) according to the manufacturer’s instructions, loaded onto a R9.4.1 flow cell. Sequencing was performed on the GridION platform running MinKNOW version 21.05.25 with live base-calling on high accuracy mode with demultiplexing enabled (Guppy version 5.0.16). Sequencing run status was monitored on board MinKNOW and run was terminated after more than 20 MB of passed base-called data was obtained per sample.

***Respiratory viral enrichment using hybridisation capture probes.*** The cDNA generated prior to whole genome amplification was used as input into the RNA Prep with Enrichment kit (Illumina). Second-strand cDNA synthesis, cDNA tagmentation, library construction, clean-up, and normalization were performed according to manufacturer’s instructions. Individual libraries were then combined in 3-plex reactions for probe hybridization. The Respiratory Viral Oligo panel v2 (Illumina) was used for probe hybridization with the final hybridization step held at 58°C overnight. Hybridized probes were then captured and washed according to manufacturer’s instructions and amplified as follows: initial denaturation 98°C for 30 s, 14 cycles of: 98°C for 10 s, 60°C for 30 s, 72°C for 30 s, and a final 72°C for 5 min. Library quantities and fragment size were determined using a Qubit 1× dsDNA HS assay and Agilent HS Tapestation and sequenced using 2 × 76-bp runs on the Illumina iSeq.

***Bioinformatic analysis of Illumina data.*** Raw sequence data were processed using an in-house quality control procedure prior to further analysis as described previously.^1,5^ De-multiplexed reads were quality trimmed using Trimmomatic v0.36 (sliding window of 4, minimum read quality score of 20, leading/trailing quality of 5 and minimum length of 36 after trimming).^6^ Briefly, reads were mapped to the reference SARS-CoV-2 genome (NCBI GenBank accession MN908947.3) using Burrows-Wheeler Aligner (BWA)-mem version 0.7.17^7^, with unmapped reads discarded. Average genome coverage was estimated by determining the number of missing bases (Ns) in each sequenced genome. For amplicon generated reads variant calling and the generation of consensus sequences was conducted using iVar^8^, with soft clipping over primer regions (version 1.2.1, min. read depth >10x, quality >20, min frequency threshold of 0.1). Single nucleotide polymorphisms (SNP) were defined based on an alternative frequency >0.75 whereas Minority allele Frequency Variants (MFV) were defined by an alternative frequency between 0.1 and 0.75 and verified using manual inspection of bam files. Variants falling in the 5’ and 3’UTR regions were excluded. Polymorphic sites that have previously been highlighted as problematic were monitored.^9^ To ensure the accuracy of variant calls only high-quality genomes with >90% genome coverage and a mean depth of >100x were included. The MFV calls were excluded in the base pair either side of the 5’ or 3'-end of indels due to potential mis-mapping. Host reads were removed from FASTQ files before being uploaded to SRA. SARS-CoV-2 lineages were inferred using Phylogenetic Assignment of Named Global Outbreak LINeages v1.2.86 (PANGO and PLEARN).^10,11^

***Bioinformatic analysis of ONT data.*** Quality control and consensus sequence were generated post run using the wf-artic workflow version 0.3.9 (<https://github.com/epi2me-labs/wf-artic>). To determine and quantify positional heterozygosity, mapping files generated by the wf-artic workflow were visualised on the Integrative Genomics Viewer^12^ version 2.8.6 and parsed using bam-readcount version 1.0.1 (<https://github.com/genome/bam-readcount>).

***Amplification bias in co-infection cases.*** Consensus and minority allele frequency variants were collated over constellations of mutations that define the SARS-CoV-2 lineages B.1.617.2 (Delta) and BA.1 (Omicron) for each genome investigated (<https://github.com/cov-lineages/constellations/blob/main/constellations/>). This included 10 unique genomic locations that define Delta (B.1.617.2) (S:T19R, S:L452R, S:P681R, ORF3a:S26L, M:I82T, ORF7a:V82A, ORF7a:T120I, N:D63G, N:R203M, N:D377Y). Mutations that co-exist in BA.1 were not included (S:G142D, S:T478K, S:D950N). In addition to 17 polymorphisms that define the dominant Omicron lineage BA.1 (orf1ab:K856R, del:6513:3, nuc:T5386G, orf1ab:A2710T, orf1ab:I3758V, nuc:C15240T, S:A67V, del:21765:6, del:21987:9, del:22194:3, nuc:22205+GAGCCAGAA, S:S371L, S:G446S, S:G496S, S:T547K, S:N856K, S:L981F, M:D3G) mutations that co-exist in B.1.617.2 where not included (S:T95I). Alternative read frequencies over each polymorphism were compared using all three sequencing techniques and between sampling timepoint of each case.

***Phylogenetic analysis of reconstructed strains.*** Representative SARS-CoV-2 genomes collected between December 2020 and 31^st^ December 2022 (n=1,300, ≥27,000-bp in length) were downloaded from the Global Initiative on Sharing All Influenza Data (GISAID)^13^ EpiCoV, using a global subsampling strategy developed by Nextstrain.^14^ Phylogenetic inference and visualisation of the 1081 high quality consensus SARS-CoV-2 FASTA sequences (GISAID, n =1076; study, n = 5). The GISAID data set included representatives of primary Delta and Omicron strains currently known to be circulating in Sydney. For Case B, the consensus genomes for the Day 0 (Omicron only, Illumina Midnight) and Day 3 (Delta dominant, Illumina RVOP to avoid the Midnight artefact) samples were used. The Delta sequence for Case A, for which all three samples were of mixed lineage, was obtained from the Day 3 sample culture. To obtain the Omicron sequence for Case A, the RVOP Illumina data from the Day 3 sample that the culture was derived from was hand-reviewed to generate a consensus containing only the Omicron-specific SNPs, minus the Delta-specific SNPs seen in the culture sequence. The resulting reconstructed Case B Omicron was then compared to the Omicron sequence from Case C, who is assumed to have caught their infection from Case B; these two sequences matched. These four sequences, as well as the downloaded sequences from GISAID, were trimmed to remove the 5’ and 3’ UTR regions and aligned with MAFFT v7.402 (FFT-NS-2, progressive method).^15^ Phylogenetic analysis was performed using the maximum likelihood approach (IQTree v1.6.7 (substitution model: GTR+F+R2) with 1,000 bootstrap replicates.^16^ The phylogenetic tree was visualised using the R package ggtree.^17^

***Statistical analysis.*** Statistical analysis of the read distribution of Delta and Omicron lineage markers was performed by Student's t-Test using R software version 4.1.2^18^ (2021-11-01) with the t.test function from the package ‘stats’. Graphs were generated using the package ‘ggplot 2’ version 3.3.5.^19^

***Human research ethics approval.*** Ethical and governance approval for the study was granted by the Western Sydney Local Health District Human Research Ethics Committee (2020/ETH02426).

**SUPPLEMENTARY RESULTS**

***Frequency of SARS-CoV-2 variant heterozygosity.*** From a total of 22,942 high quality genomes (minimum 90% coverage of the SARS-CoV-2 genome) sequenced between 25 February 2020 and 20 January 2022, only 10 (0.04%) had 30 or more heterozygous sites (min 0, median 1, mean 1.28, max 77, 80^th^ percentile 2, 99^th^ percentile 6, 99.9^th^ percentile 17) (Figure S2). Sequences with ≥ 10 heterozygous sites trigger a sequence quality review. A total of 33 heterozygous sites were initially detected in the genome of Case A (Day 2) and 46 heterozygous sites from Case B (Day 3). Failure to pass this bioinformatic quality metric resulted in a complete repeat of the SARS-CoV-2 genomic workflow for these samples, including re-extraction of the original primary specimen.

***Patient cohort details***. Three specimens were collected from each case and are summarised in Table S1. The low SARS-CoV-2 viral load in the index specimen from each case resulted in a SARS-CoV-2 genome only being generated from the Illumina sequencing methodology, (Case B) and an incomplete genome from Case A (90% coverage).

***Investigation of SARS-CoV-2 amplification bias in Omicron and Delta co-infection.*** Amplification biases noted in ONT and Illumina data are caused by lineage specific mutations impacting the efficiency of amplification primers. These biases resulted in increased variation in the frequency of lineage defining mutations, particularly when comparing amplification-based SARS-CoV-2 enrichment to probe capture-based methodologies (Figure S3 and S4). Amplification of Omicron subpopulations is reduced in the region encompassing Midnight amplicon 28 (nt 27808 – 28985). This is due to a mismatch at the 3’end of forward primer 28 (TTTGTGCTTTTTAGCCTTTCTG**C**(t-Omicron)T. Mutations in the last 5bp of primer sequences significantly impact the efficiency of amplification, therefore Delta populations were preferentially amplified over lineage markers ORF7a:V82A, ORF7a:T120I (Table S1). Similar biases were noted in Omicron lineage markers (S:N856K, S:L981F) due to a primer mismatch in forward primer 24 (GCTGAA**C**(t-Omicron)ATGTCAACAACTC). Although not a lineage defining mutation, a 17-bp deletion in ORF7a predominated in Delta strains circulating in Australia between July and December 2021. This deletion was not detected in the Delta-predominant consensus genomes of Case B, Day 11 in genomes produced using Midnight primer amplification, but was present in the genomes produced using RVOP. A Delta-specific mutation in the Midnight reverse primer 27 at the 3’end of the primer was detected, likely causing preferential amplification of the Omicron subpopulation in this region, encompassing Midnight amplicon 27. A similar phenomenon was recently reported for a collection of Delta samples with minor Omicron contamination sequenced using ARTIC V3.^20^

**SUPPLEMENTARY TABLES AND FIGURES**

**Supplementary Table S1**. SARS-CoV-2 yield and sequencing metrics for co-infection cases.

**Supplementary Table S2.** Acknowledgement of SARS-CoV-2 genomes used in the study, generously contributed by international laboratories on the GISAID repository.

**Supplementary Figure S1.** Bioinformatics workflow, highlighting critical quality performance parameters used.

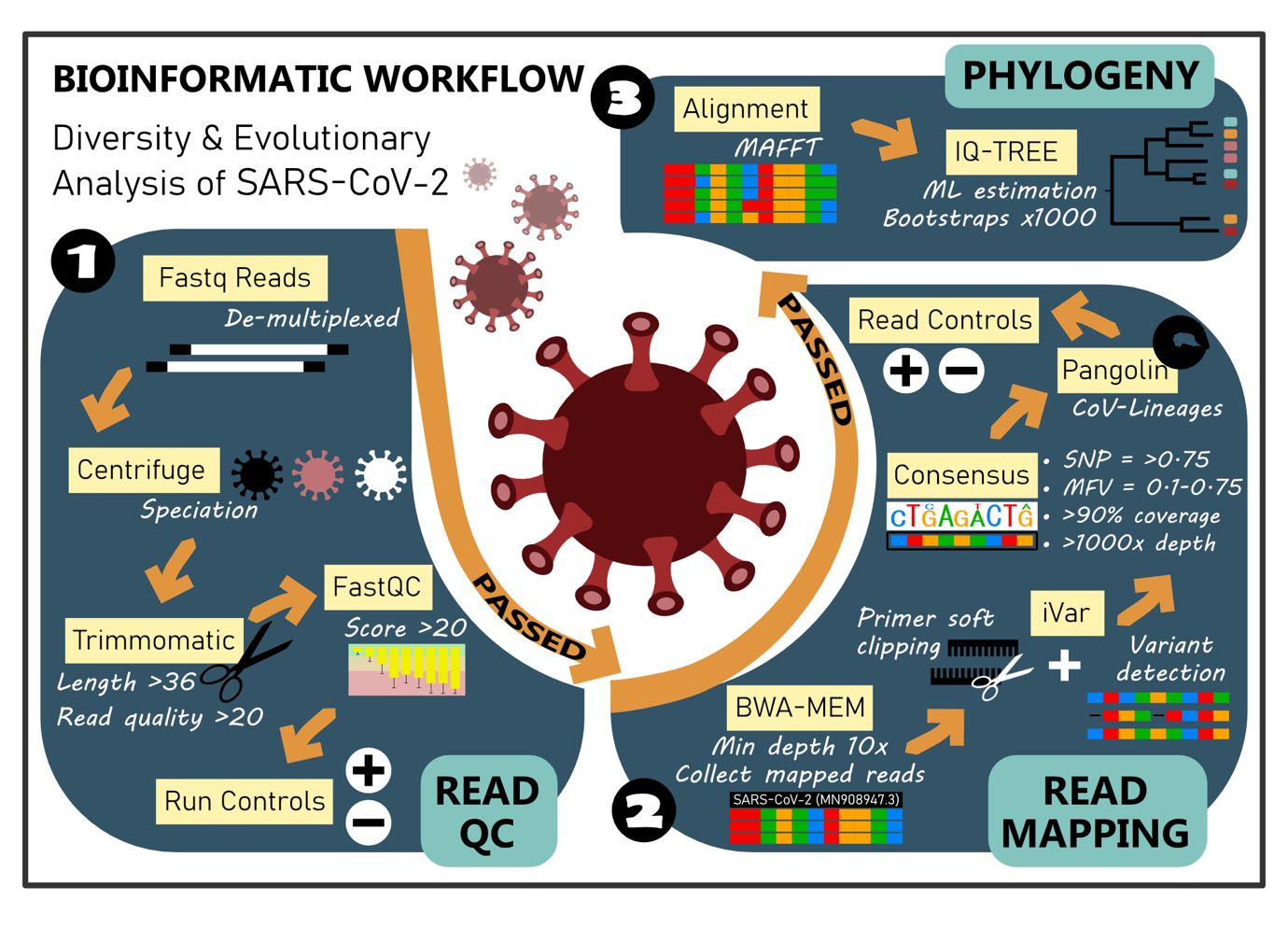

**Supplementary Figure S2.** Log-scale histogram of heterozygosity in 22,2942 high quality (≥90% SARS-CoV-2 genome coverage) SARS-CoV-2 genomes produced from NSW, Australia. Larger dark blue dots indicate the genomes investigated in this study, the regression is depicted by the blue line, with the grey area showing the 95% confidence intervals.

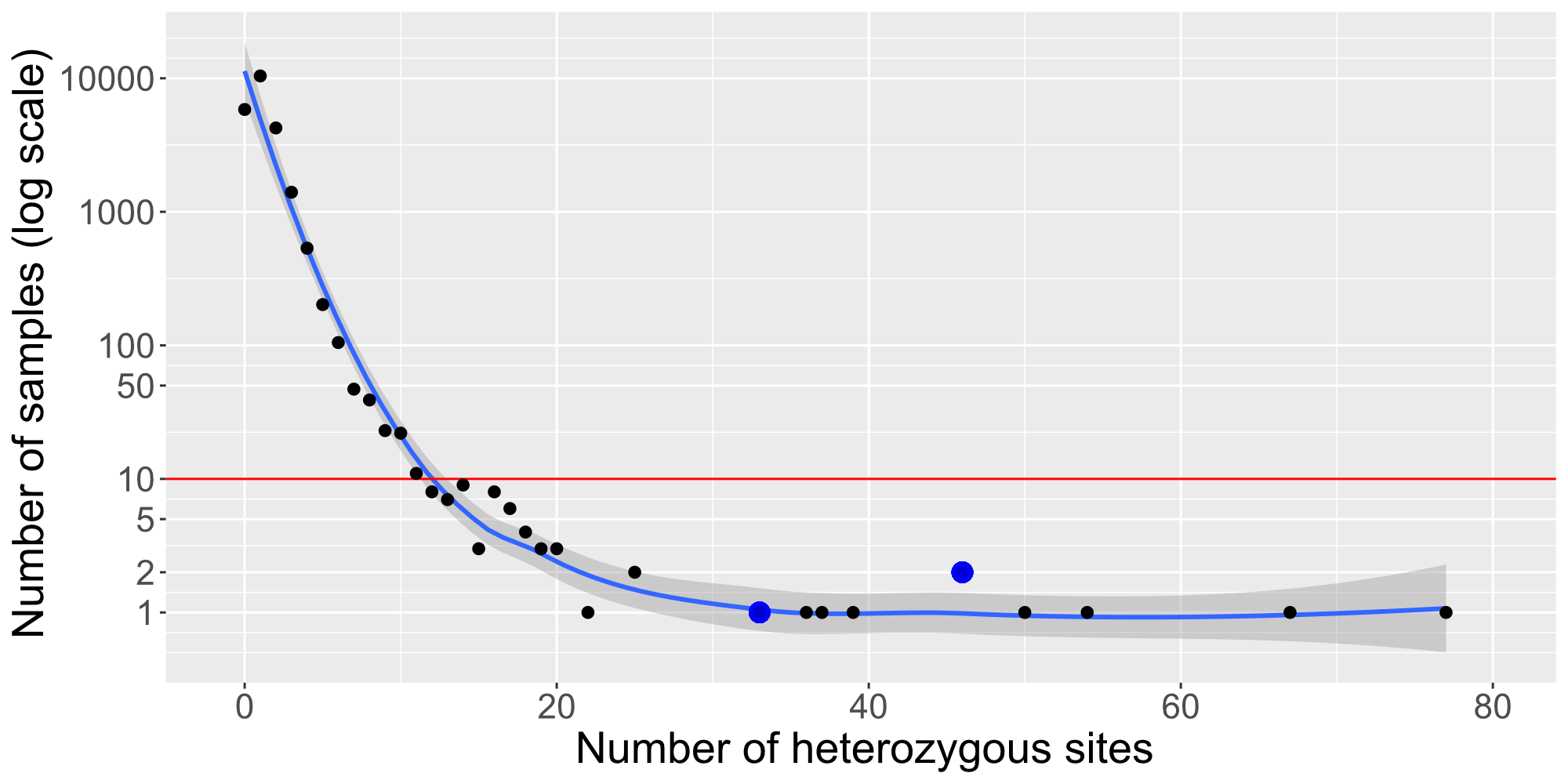

**Supplementary Figure S3.** Comparison of sensitivity of different sequencing methods and technologies to detect SARS-CoV-2 co-infection with the Delta and Omicron lineages. Average population frequency of the Omicron and Delta-lineage defining mutations in clinical samples by different sequencing method. Segments in grey represent differences in Omicron and Delta mutational frequencies.

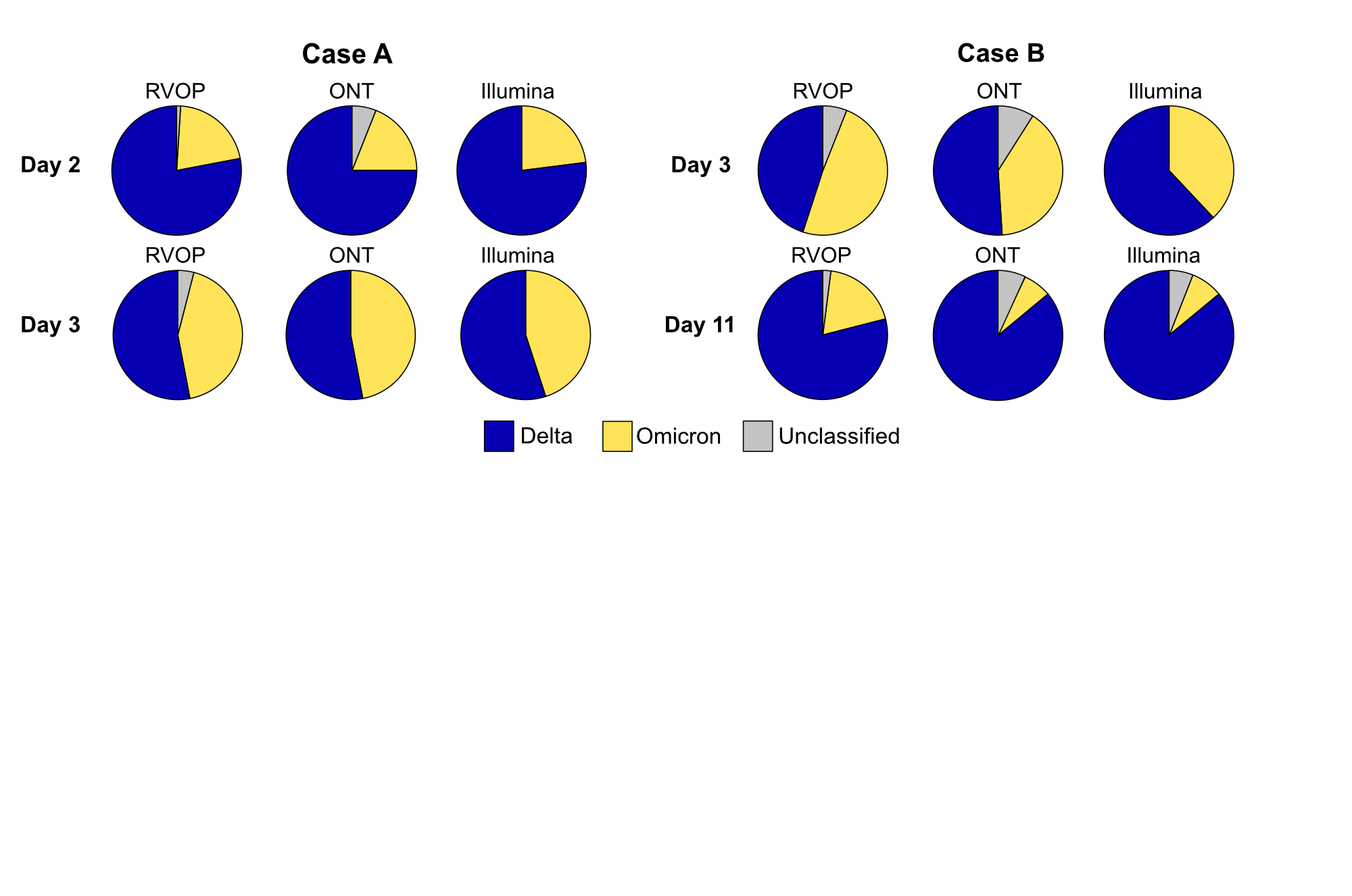

**Supplementary Figure S4.** Read frequency distribution of Delta and Omicron lineage markers using three SARS-CoV-2 sequencing methods. Both Delta and Omicron lineages markers were detected **Case A** on Day 2, with Delta being the dominant lineage. On Day 3, the difference was reduced, with no difference in the proportion of Delta / Omicron detected by Illumina or ONT. RVOP still detected a significant difference in the proportion of Delta and Omicron lineage markers.  For **Case B** on Day 3 - there was more Delta than Omicron lineage detected by Illumina sequencing, but no difference in the proportion of Delta/Omicron detected by ONT or RVOP. By Day 11 the proportion of Delta/Omicron has increased as detected by all three-sequencing methods NS (not significant) p > 0.05, * p <= 0.05, *** p <= 0.001. Sequencing methods: ONT - Oxford Nanopore Technologies sequencing, Illumina – Illumina short read sequencing, RVOP - Respiratory Virus Oligo Panel.

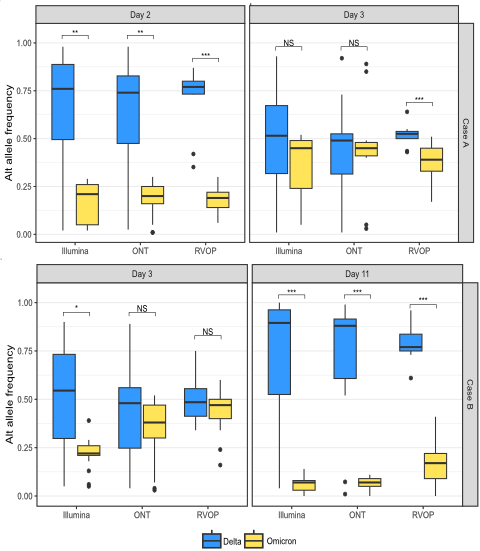

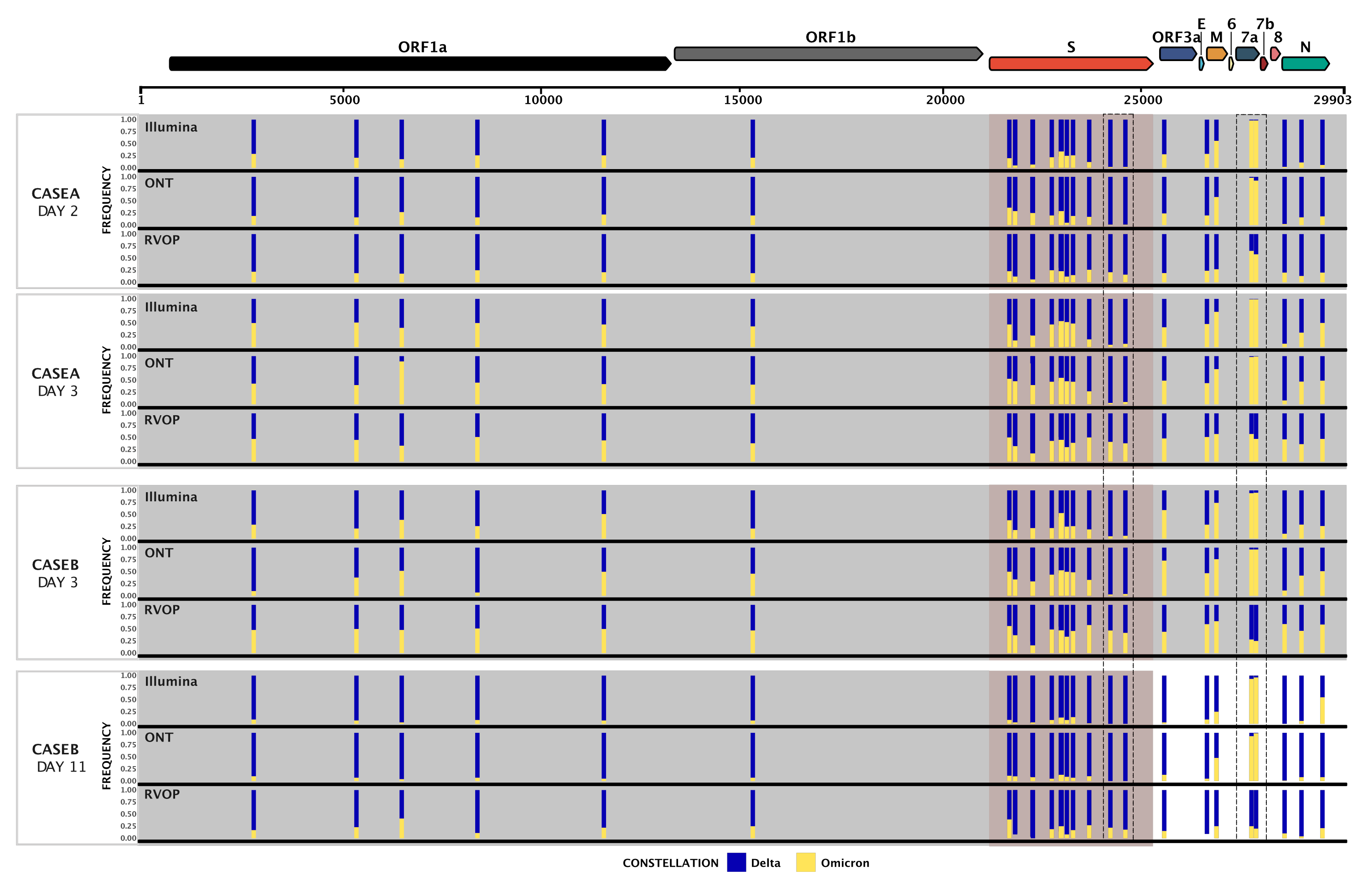

**Supplementary Figure S5.** Genome-wide view of SARS-CoV-2 with the variant frequency of Delta and Omicron lineage-defining polymorphisms in specimens sequenced using three SARS-CoV-2 sequencing methodologies. A total of 27 polymorphism defining Delta and Omicron lineages are shown in relation to the annotated SARS-CoV-2 genome. The frequency of sequencing reads encoding each mutation is shown by histograms highlighting the constellation of mutations defining each lineage. Blue bars demonstrate the frequency of mutations defining the Delta lineage and yellow bars the Omicron lineage. Due to the close genomic location of lineage defining mutations in the spike region some bars are overlapping. Read frequencies are collected from ONT, Illumina and RVOP data and although there is high concordance between sequencing methods and technologies, the dashed line inserts indicate regions of amplification bias that preferentially amplify Delta or Omicron in ONT and Illumina sequencing results, in contrast to RVOP data where lineage defining mutational frequencies remain more consistent.
